# Harmonizing Healthy Cohorts to Support Multicenter Studies on Migraine Classification using Brain MRI Data

**DOI:** 10.1101/2023.06.26.23291909

**Authors:** Hyunsoo Yoon, Todd J. Schwedt, Catherine D. Chong, Oyekanmi Olatunde, Teresa Wu

**Author notes:** Corresponding Author: Teresa Wu, Professor, School of Computing and Augmented Intelligence, Arizona State University, Adjunct Professor of Radiology, College of Medicine, Mayo Clinic, Founding Director, ASU-Mayo Center for Innovative Imaging. **Data Availability Statement:** Data from the study sponsored by the United States Department of Defense (DOD) will be made available through the Federal Interagency Traumatic Brain Injury Research (FITBIR) Informatics System in accordance with the rules and regulations of the DOD. Patient consent for the NIH-sponsored study and for the Mayo-funded study did not include a data sharing agreement.

## Abstract

Multicenter and multi-scanner imaging studies might be needed to provide sample sizes large enough for developing accurate predictive models. However, multicenter studies, which likely include confounding factors due to subtle differences in research participant characteristics, MRI scanners, and imaging acquisition protocols, might not yield generalizable machine learning models, that is, models developed using one dataset may not be applicable to a different dataset. The generalizability of classification models is key for multi-scanner and multicenter studies, and for providing reproducible results. This study developed a data harmonization strategy to identify healthy controls with similar (homogenous) characteristics from multicenter studies to validate the generalization of machine-learning techniques for classifying individual migraine patients and healthy controls using brain MRI data. The Maximum Mean Discrepancy (MMD) was used to compare the two datasets represented in Geodesic Flow Kernel (GFK) space, capturing the data variabilities for identifying a “healthy core”. A set of homogeneous healthy controls can assist in overcoming some of the unwanted heterogeneity and allow for the development of classification models that have high accuracy when applied to new datasets. Extensive experimental results show the utilization of a healthy core. One dataset consists of 120 individuals (66 with migraine and 54 healthy controls) and another dataset consists of 76 (34 with migraine and 42 healthy controls) individuals. A homogeneous dataset derived from a cohort of healthy controls improves the performance of classification models by about 25% accuracy improvements for both episodic and chronic migraineurs.

**Highlights:**

- The harmonization method was established by Healthy Core Construction.
- The inclusion of a healthy core addresses intrinsic heterogeneity that exists within a healthy control cohort and in multicenter studies.
- The utilization of a healthy core can increase the accuracy and generalizability of brain imaging-based classification models.
- The proposed harmonization method offers flexible utilities for multicenter studies.

## 1. Introduction

Neuroimaging studies have shown structural and functional alterations in brain regions involved in sensory-discriminative, affective, and cognitive pain processing, pain modulation, and multisensory integration in patients with migraine [Eck, J. et al. 2011, Lo Buono et al. 2017, Meylakh, N. et al. 2018, Mu, J. et al. 2020, Wei H. et al. 2020, Scheepens, D. S. 2020, Ashina, S. 2021]. Machine learning models trained to automatically classify individuals with migraine from healthy controls (HCs) using functional magnetic resonance imaging (fMRI) and structural MRI data have shown good utility for discriminating those with migraine from HCs. Yet, the generalization of the algorithms might be poor; that is, the classification model developed using one dataset may not perform as accurately on a separate dataset. The generalizability of classification models is key for multi-scanner and multicenter studies, and for providing reproducible results. Harmonization in clinical research has been focusing on defining, reviewing, and standardizing common data elements [Firnkorn, D et al. 2015, Yu, M. et al. (2018), Ma, D. et al. (2019), Chen, A. A. et al. (2022)]. This is known as data curation - a strategy to combine the data from different sources to improve the generalizability of classification models. To be specific, this harmonization strategy is the explicit removal of the site or cohort-related effects in data from multiple sources. This harmonization strategy is desirable for improving the generalizability of classification models. Yet, from a data-driven aspect, the data from different sources may inherently exhibit different characteristics due to several reasons, such as differences in patient populations from which research participants are identified and the methods by which data are acquired. In machine learning research, extensive efforts are dedicated to transforming the data from different sources into a common feature space for evaluating similarities from multiple sources or adapting them thoroughly. This is also known as Domain Adaptation, a well-studied field [Tzeng, et al. 2017, Csurka et al. 2017, Tang, et al. 2020].

Domain adaptation leverages labeled data in one or more related source domains, to build a machine learning model for unseen or unlabeled data in a target domain. These adaptation methods aim to discover a shared feature representation for minimizing distribution differences while preserving the important properties of original datasets. There are several types of domain adaptation approaches, such as re-weighting, parameter adaptation, feature augmentation, and feature transformation [Yao, Y et al. 2010, Gong, B 2012, Matasci, G 2015, Tzeng, et al. 2017, Csurka et al. 2017, Tang, et al. 2020]. After adaptation, a standard machine learning approach is used that assumes the test data are drawn from a similar distribution as the training data. As noted, most domain adaptation methods are designed to transform the data (both HCs and patients) together into a common space. Careful consideration of this approach reveals a potential concern, that is, it ignores the fact that there may exist heterogeneity even within the HC cohorts. As a result, transforming and aligning the whole dataset may not work as well as expected. In this research, we propose a new concept termed “healthy core”: that transforms and aligns imaging data from HCs using their brain MRI data. Specifically, this study aims to 1) identify HCs from different sources to construct a healthy core; 2) develop and test classification models based on medical imaging that utilizes the healthy core in model training, validation, and testing across the datasets. We hypothesized that, compared to classification models developed without the use of a healthy core, imaging-based classification models for migraine developed with the use of a healthy core would have higher classification accuracy when applied to an unseen dataset.

## 2. Method

### 2.1. Approvals and Consent

All studies were approved by the Mayo Clinic and Washington University School of Medicine in St. Louis Institutional Review Boards. All participants provided written consent prior to study participation. Individuals with migraine were identified from the headache clinics at both institutions. HCs and individuals with migraine were enrolled from a database of research volunteers, and via community outreach.

### 2.2. Study Participants

Diagnoses of episodic migraine (EM) and chronic migraine (CM) were assigned using the most recent version of the International Classification of Headache Disorders available at the time of enrollment [ICHD-3 beta 2013]. Migraine diagnoses were assigned by a headache specialist (TS). HCs were excluded if they had a history of migraine, but occasional tension-type headaches (< 3 tension-type headaches per month) were allowed. Participants were male or female adults between the ages of 18-65. Those with contraindications to MRI, acute pain conditions other than migraine, neurological disorders other than migraine, women who were pregnant, and individuals with abnormal brain MRI findings according to usual clinical interpretation were excluded. Participants were recruited for the study between 2012 and 2021.

### 2.3. Data Collection

<u>Dataset 1 (DS1):</u> The DS1 cohort, a total of 120 individuals (66 with migraine and 54 HCs) includes some patients scanned at Mayo Clinic Arizona and other patients scanned at Washington University School of Medicine in St. Louis.

Dataset 2 (DS2): The DS2 cohort, a total of 76 (34 with migraine and 42 HCs), consists of data from individuals scanned only at Mayo Clinic Arizona.

Data collected from all study participants included age, sex, Beck Depression Inventory-II (BDI-II) scores, and State-Trait Anxiety Inventory (STAI) scores, Additional data collected from migraine participants included headache frequency, number of years lived with headache, and Migraine Disability Assessment (MIDAS) scores.

Study participants were imaged on one of two Siemens (Erlangen, Germany) scanners, each at a different institution. Scanners at both institutions differed in scanner model (Magnetom vs Trio), use of headcoil (12-channel vs 20-channel), and T1-weighted and T2-weighted acquisition parameters, described in detail below.

Washington University: MAGNETOM Trio 3T scanner using a 12-channel head matrix coil

T1-weighted images: TE=3.16 ms, TR=2.4 s, 1×1×1 mm voxels, 256×256 mm field of view (FOV), acquisition matrix 256 x 256. T2-weighted images: TE=88 ms, TR=6280 ms, 1×1×4 mm voxels, 256×256 mm field of view, acquisition matrix 256 x 256.

Mayo Clinic: MAGNETOM Skyra 3T scanner using a 20-channel head matrix coil.T1-weighted images: TE=3.03 ms; TR=2.4 s; 1×1×1.25 mm voxels; 256×256 mm field of view, acquisition matrix 256 x 256. T2-weighted images: TE=84 ms; TR=6800 ms; 1×1×4 mm voxels; 256×256mm FOV, acquisition matrix 256 x 256).

T1 images were used for ruling out gross structural abnormalities.

Data included in this analysis have been utilized in prior analyses [Schwedt, T.J., 2015 and Chong, C. D. 2017]. T1-weighted image processing was performed using the automated FreeSurfer image analysis suite (version 5.3, http://surfer.nmr.mgh.harvard.edu/) available at the time of imaging. Image processing included skull stripping, automated Talairach transformation, segmentation of subcortical gray and white matter, intensity normalization, gray-white mater boundary tessellation, and surface deformation[Dale, A. M (1999), Fischl, B. (2001), Fischl, B. (2002), Ségonne, F. (2004)]. Data output measures from the automated ‘recon-all’ processing stream included regional cortical volume, cortical surface area, and cortical thickness over the left and right hemispheres.

### 2.4. Evaluation of Dataset differences between DS1 and DS2

For both datasets, measurements of cortical thickness, cortical surface area, and cortical volume for 68 regions were derived resulting in 204 measures per subject. Image features of HCs on cortical thickness, cortical surface area, and volumes in DS1 and DS2 were compared separately to identify the potential cohort difference. The comparison was performed for the means of features, the variances of features, and feature correlation structures. The cohort-wise means/variances of each feature were compared using a two-sample t-test/F-test. False Discovery Rate (FDR) was used to correct for multiple comparisons. The cohort-wise correlation matrices for the volume, area, and thickness features were compared using the two-sample Hotelling’s T2 (the multivariate extension of the common two-group Student’s t-test) and Multivariate Analysis of Covariance. In order to avoid the confounding of multiple cohort differences, the mean and variance differences were removed from each cohort to identify the remaining differences.

### 2.5. Constructing the Healthy Core

Differences in imaging acquisition protocols, scanners, and even small differences in populations from which research participants are enrolled can lead to cohort effects. Therefore, simply combining all HCs from different studies/sites may not be optimal due to cohort effects. In order to address this issue, here we propose a new design strategy to discover the homogenous HCs from different datasets as referenceable HCs (see Step 2 in Figure 1). Our proposed approach contains two key concepts: 1) measure the similarity between HCs from different datasets, and 2) explore feature representations of the dataset to assess the similarities. First, the proposed method utilizes the Maximum Mean Discrepancy (MMD) [Gretton, A et al. 2012, Cui, P et al. 2020] which is a kernel-based statistic used to determine whether two distributions are similar. MMD can be considered as a criterion to determine whether subset samples from multiple datasets are homogenous [Gretton, A et al. 2012, Long, et al. 2015, Yan, et al. 2017, Epstein, et al. 2019, Zhang, et al. 2020, Kirchler, et al 2020]. However, the MMD is a point estimate-based statistic which may be too strict since the original datasets might inherently exhibit different characteristics. To capture the inherent variabilities, we hypothesize that Geodesic Flow Kernel (GFK), which utilizes geometric manifolds termed Grassmann, may help. GFK has been widely used in the domain adaptation research field because it does not focus only on the feature representations of the two datasets (snapshot of each dataset), it constructs the transformation over the Grassmann space of the two datasets with a scale parameter. As a result, it has the potential to identify the subset samples from each dataset, for example, using MMD, as the homogenous core.

**Figure 1.**
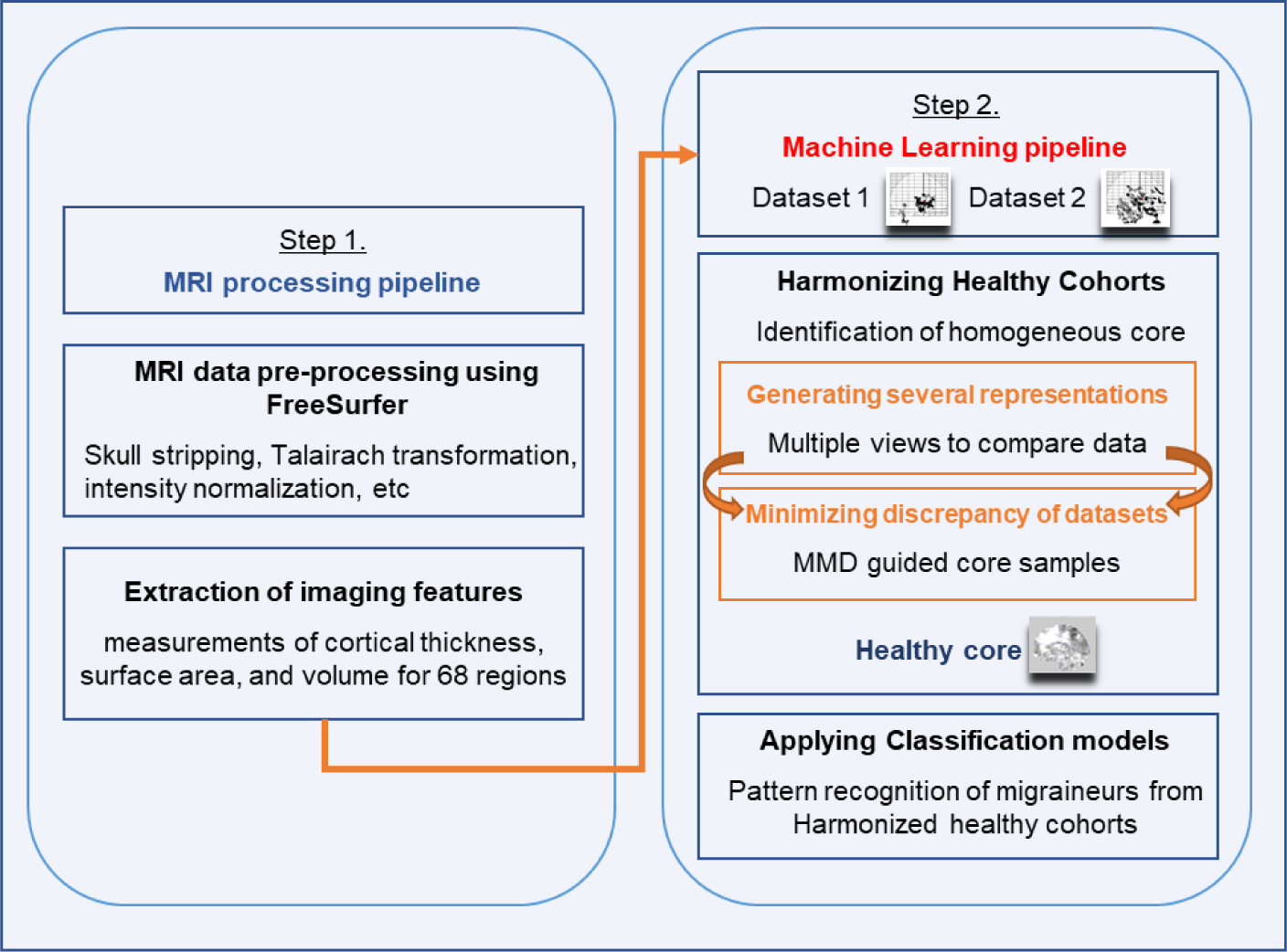
The overall workflow for Healthy Core Construction

We hypothesize that using a healthy core, a homogenous subset of HCs from different sources, can help improve the generalization of the machine learning model for classifying migraine since it addresses the heterogeneity of the HCs within and across the datasets; and sets the bridges between the datasets. To comprehensively assess the clinical utility of the proposed method, we designed three sets of experiments: (1) evaluating the performance of migraine classification models within a single dataset; (2) evaluating the performance of migraine classification models developed using one dataset and tested on a second dataset; and (3) evaluating the performance of migraine classification models with and without using healthy core samples for classifying individuals with CM and those with EM from a different dataset. In the training of the classification models, cross-validation has been applied. To get reliable results for the relatively small sample sizes, experiments are repeated under five different random scenarios (cross-validated) to build classifiers and report average performances. Since our last experiment focuses on migraine only, we report the Area Under the Curve (AUC) for the first two experiments and the accuracy of identifying migraine in the last experiment.

**Figure 2.**
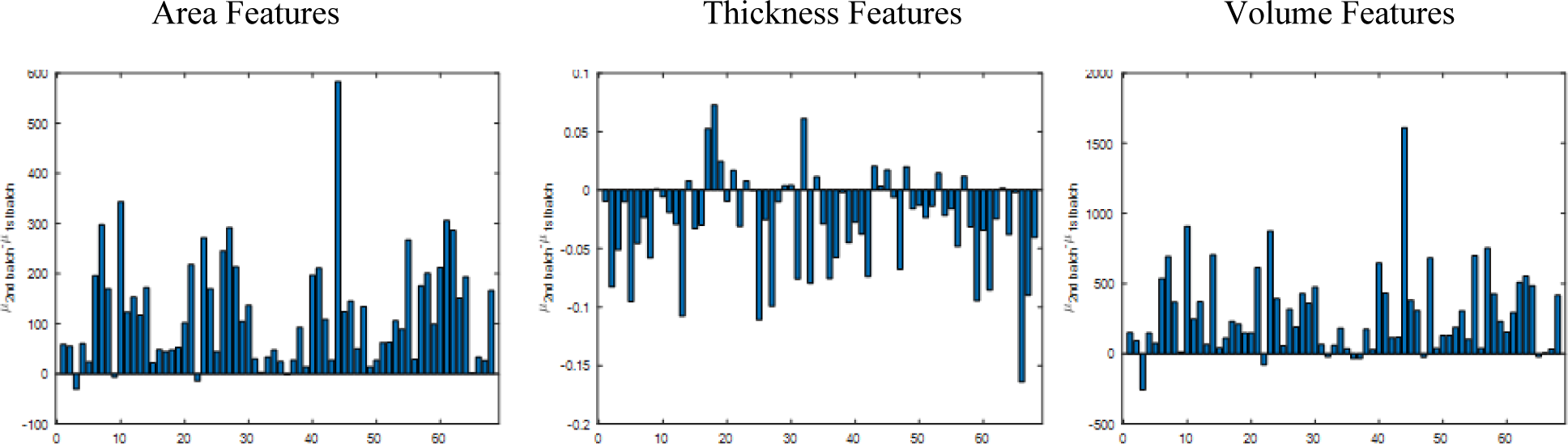
Univariate Analysis for Mean Difference Scale

## 3. Results

DS1 consisted of 120 subjects (Age: 36 ± 11, female:91, male:29, Amongst those with migraine, an average headache frequency was:8.91 ± 6.25 days per month), including 54 HC, 51 EM patients, and 15 CM patients. DS2 consisted of 76 subjects (Age: 38 ± 10, female:41, male:35, Average headache frequency for those with migraine: 17.97 ± 8.08 days per month), including 42 HCs, 8 EM patients, and 26 CM patients. The overall sample numbers in this study are shown in Table 1 and the proportion of individuals with aura is described in Table 2.

**Table 1.** Overall sample numbers for HC, EM, and CM

|  | HC | EM | CM |
| --- | --- | --- | --- |
| DS1 (120 subjects) | 54 | 51 | 15 |
| DS2 (76 subjects) | 42 | 8 | 26 |

**Table 2.** Proportion of individuals with aura

|  | DS1 (66 migraineurs) |  | DS2 (34 migraineurs) |  |
| --- | --- | --- | --- | --- |
|  | EM | CM | EM | CM |
| Aura | 22 | 6 | 3 | 15 |
| No Aura | 29 | 8 | 5 | 11 |
| Unknown | 0 | 1 | 0 | 0 |

### 3.1. Dataset (Cohort) Differences

First, the mean of area, thickness, and volume features are significantly different between the HCs in the two datasets with a *p*-value<0.01 before the False Discovery Rate (FDR) correction. Further, three imaging features among those 41 significant features were identified as significantly different between two cohorts by the two-sample t-test and Kolmogorov-Smirnov test after FDR correction (Table 3).

**Table 3.** The difference in the mean of features between DS1 and DS2

|  | Two-sample t-test | Two-sample Kolmogorov-Smirnov |
| --- | --- | --- |
| Before FDR | 41 | 43 |
| After FDR | 12 | 3 |

Second, the variances of the features were tested. Table 4 shows that the variance for each feature is not significantly different between two cohorts under both tests after FDR correction.

**Table 4.** The difference in the variances of features between DS1 and DS2

|  | Two-sample *F*-test for equal variances | Ansari-Bradley Test for Equal Variances |
| --- | --- | --- |
| Before FDR | 8 | 9 |
| After FDR | 0 | 0 |

Furthermore, we investigated the possibility that significant group differences in sex distribution (*p*-value=0.01) contributed to the cohort difference.

We randomly selected the sex-balanced sub samples from both cohorts. After making the balanced groups, the mean and variance of features (volume, area, and thickness) were tested using the t-test and KS test for mean, F-test and Ansari-Bradley test for variance whether the dataset difference exists. Since we selected a random subset to test, this test procedure was repeated 10,000 times to avoid a sampling-dependent conclusion. It was concluded that there is a significant mean difference and a marginal variance difference in each feature (volume, area, and thickness), but sex distribution was not a significant factor in the dataset difference.

Table 5 shows the average mean difference under several scenarios such as DS1 vs DS1, DS2 vs DS2, Random splits, and DS1 vs DS2. DS1 vs DS1 illustrates 21 randomly selected samples from the DS1 dataset and 21 non-overlapped samples from DS1. Note 21 samples are the number of consistent and balanced samples across four scenarios. The random splits mean 21 randomly chosen subsets from the combined dataset (DS1 and DS2) and another non-overlapped 21 samples. The number in the table shows the mean features per each subset and their differences. Because of the randomness in the sampling, we repeat this task 100 times and report the averaged values.

**Table 5.** Average Mean Difference under Random Subsets within Dataset

| Avg. \|DS1-DS2\| | DS1 vs DS1 | DS2 vs DS2 | Random splits | DS1 vs DS2 |
| --- | --- | --- | --- | --- |
| Area (mm<sup>2</sup>) | 82.12 | 77.86 | 80.75 | 102.27 |
| Thickness (mm) | 0.0416 | 0.0458 | 0.0436 | 0.0481 |
| Volume (mm<sup>3</sup>) | 253.74 | 257.07 | 256.01 | 275.58 |

Table 5 confirms there is more of a difference between the two datasets (DS1 vs DS2) compared to sample subsets from within a dataset (e.g., DS1 vs DS1). The test results indicate that DS1 and DS2 are not compatible after mean and variance adjustments. Simple cohort-wise adjustments still have limited clinical applicability. This finding motivated us to develop new methods to construct a “healthy core” that consists of HC with similar characteristics from DS1 and DS2.

### 3.2. Experiment I: evaluating the performance of different classification models within a single dataset

The goal is to demonstrate the accuracy of machine learning models in classifying migraine within a single dataset as shown in table 6. In this experiment, we built four binary classifiers (regularized logistic regression, support vector machine (SVM), random forest, and XGBoost) to identify HC vs. CM, or HC vs EM. For each dataset (DS1 or DS2), we split the data into training (∼60%), validation (∼20%), and testing (∼20%).

**Table 6.** Single Dataset Experiments

|  | Dataset | Training | Validation | Test |
| --- | --- | --- | --- | --- |
| CM | DS 1 | HC (n=34) vs. CM (n=9) | HC (n=9) vs. CM (n=3) | HC (n=11) vs. CM (n=3) |
|  | DS 2 | HC (n=27) vs. CM (n=17) | HC (n=7) vs. CM (n=4) | HC (n=8) vs. CM (n=5) |
| EM | DS 1 | HC (n=34) vs. EM (n=31) | HC (n=9) vs. EM (n=10) | HC (n=11) vs. EM (n=10) |
|  | DS 2 | HC (n=27) vs. EM (n=4) | HC (n=7) vs. EM (n=2) | HC (n=8) vs. EM (n=2) |

Four different models were applied to build the classifiers. The average AUC of the four models for HC vs CM was 0.7533 for the DS1 test dataset and 0.6465 for the DS2 test dataset as seen in Table 7A. The average AUC of the four models for HC vs EM was 0.6965 for the DS1 test dataset and 0.6135 for the DS2 test dataset as seen in Table 7B. We conclude that the machine learning models provide good performance during training, but that the performance deteriorates when using test data. This deterioration in accuracy is expected and the model performance may still be considered satisfactory.

**Table 7A.** AUC of Four Classification Models in Single Dataset Experiments for CM

|  | Training on DS1 | Test on DS1 | Training on DS2 | Test on DS2 |
| --- | --- | --- | --- | --- |
| Regularized Logistic Reg. | 0.9600 | 0.7360 | 0.9820 | 0.6380 |
| SVM | 0.8720 | 0.6700 | 0.9800 | 0.5900 |
| Random Forest | 0.9320 | 0.7700 | 0.9180 | 0.6560 |
| XGBoost | 0.9480 | 0.8370 | 0.9040 | 0.7020 |
| Average |  | 0.7533 |  | 0.6465 |

**Table 7B.** AUC of Four Classification Models in Single Dataset Experiments for EM

|  | Training on DS1 | Test on DS1 | Training on DS2 | Test on DS2 |
| --- | --- | --- | --- | --- |
| Regularized Logistic Reg. | 0.8960 | 0.6660 | 0.8920 | 0.5760 |
| SVM | 0.9480 | 0.6740 | 0.9500 | 0.6060 |
| Random Forest | 0.9960 | 0.7400 | 0.9420 | 0.6280 |
| XGBoost | 0.8440 | 0.7060 | 0.9320 | 0.6440 |
| Average |  | 0.6965 |  | 0.6135 |

### 3.3. Experiment II: evaluating the performance of the classification models developed on one dataset and tested on a second dataset

The goal is to assess the generalizability of machine learning models across datasets as shown in Table 8. Like Experiment I, here we tested on HC vs. EM, or HC vs. CM. Since one dataset can be fully utilized in training and validating with testing to be conducted in the second dataset, we split the data into training (∼80%) and validation (∼20%) for DS1, or DS2 respectively.

**Table 8.** Cross-Dataset Experiments

| Datasets | Training | Validation | Test |
| --- | --- | --- | --- |
| CM | DS1: | DS1: | DS2: |
| DS1> DS2 | HC (n=43) vs. CM (n=12) | HC (n=11) vs. CM (n=3) | HC (n=42) vs. CM (n=26) |
| CM | DS2: | DS2: | DS1: |
| DS2 > DS1 | HC (n=33) vs. CM (n=21) | HC (n=9) vs. CM (n=5) | HC (n=54) vs. CM (n=15) |
| EM | DS1: | DS1: | DS2: |
| DS1 > DS2 | HC (n=43) vs. EM (n=41) | HC (n=11) vs. EM (n=10) | HC (n=42) vs. EM (n=8) |
| EM | DS2: | DS2: | DS1: |
| DS2 > DS1 | HC (n=33) vs. EM (n=6) | HC (n=9) vs. EM (n=2) | HC (n=54) vs. EM (n=51) |

As seen in Table 9A for HC vs CM, all four trained classifiers did not perform well in this cross-dataset experiment. The average AUC on models trained on DS1 dropped from 0.7533 (testing on DS1) to 0.5245 (testing on DS2), and the average AUC on models trained on DS2 dropped from 0.6465 (testing on DS2) to 0.6230 (testing on DS1). We can also observe similar patterns for HC vs EM in the cross-dataset experiment as seen in Table 9B. The average AUC on models trained on DS1 dropped from 0.6965 (testing on DS1) to 0.5781 (testing on DS2), and the average AUC on models trained on DS2 dropped from 0.6135 (testing on DS2) to 0.5963 (testing on DS1). This result demonstrates the existence of dataset discrepancies that need to be cautiously handled when building classification models.

**Table 9A.**
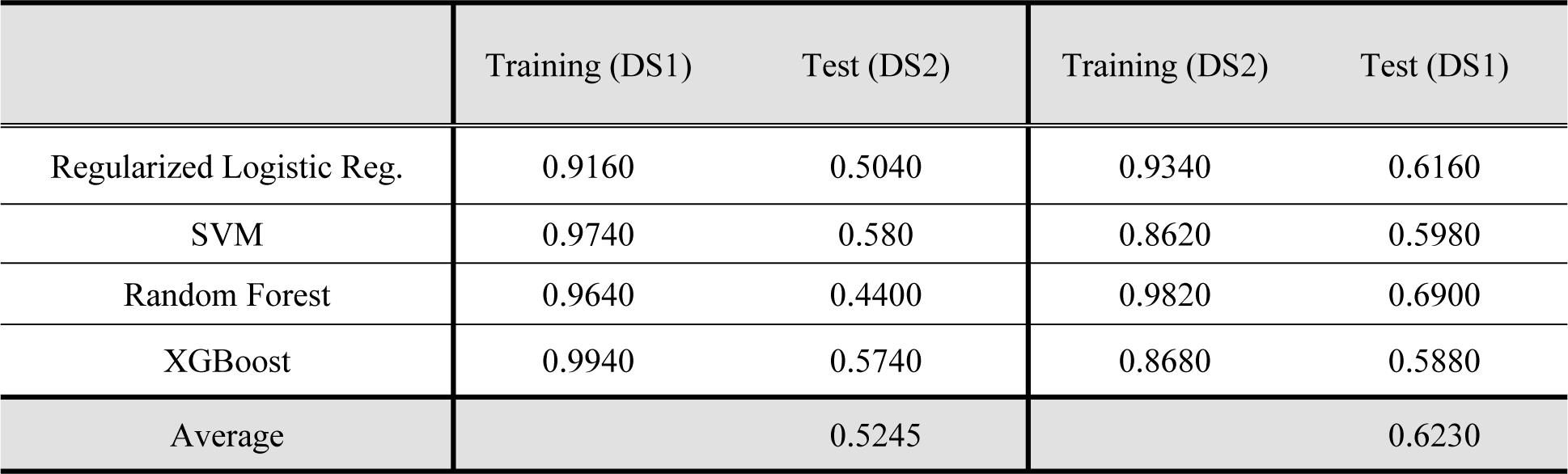
AUC of Four Classification Models for HC vs CM in Cross-Dataset Experiments

**Table 9B.** AUC of Four Classification Models for HC vs EM in Cross-Dataset Experiments

|  | Training (DS1) | Test (DS2) | Training (DS2) | Test (DS1) |
| --- | --- | --- | --- | --- |
| Regularized Logistic Reg. | 0.8475 | 0.6375 | 0.9100 | 0.5900 |
| SVM | 0.7850 | 0.6050 | 0.9725 | 0.5825 |
| Random Forest | 0.9600 | 0.5325 | 0.9675 | 0.5950 |
| XGBoost | 0.8225 | 0.5375 | 0.9075 | 0.6175 |
| Average |  | 0.5781 |  | 0.5963 |

### 3.4. Experiment III: evaluating the performance of classification models with and without using Healthy Core

The goal is to assess the applicability of machine learning models developed in one dataset including participants with migraine and the healthy core to classify individuals with migraine from a separate dataset. The overall sample numbers in Cross-dataset Experiments with Healthy Core are shown in Table 10.

**Table 10.** Cross-dataset Experiments with Healthy Core

| Datasets | Training | Validation | Test |
| --- | --- | --- | --- |
| CM | Healthy Core (n = 28) vs. | Healthy Core (n=28) vs. | CM from DS2 |
| DS1 → DS2 | CM from DS1 (n=12) | CM from DS1 (n=3) | (n=26) |
| CM | Healthy Core (n=28) vs. | Healthy Core (n=28) vs. | CM from DS1 |
| DS2 → DS1 | CM from DS2 (n=21) | CM from DS2 (n=5) | (n=15) |
| EM | Healthy Core (n = 28) vs. | Healthy Core (n=28) vs. | EM from DS2 |
| DS1 → DS2 | EM from DS1 (n=41) | EM from DS1 (n=10) | (n=8) |
| EM | Healthy Core (n = 28) vs. | Healthy Core (n=28) vs. | EM from DS1 |
| DS2 → DS1 | EM from DS2 (n=6) | EM from DS2 (n=2) | (n=51) |

Without using the healthy core, the four trained classification models do not have generalization accuracy to classify CM (test accuracy for DS2: 0.4760 – about 12 out of 26 CM, and test accuracy for DS1: 0.5229– about 8 out of 15) or EM (test accuracy for DS2: 0.5434 – about 4 out of 8 and test accuracy for DS1: 0.4903 – about 25 out of 51). Unlike typical approaches, the proposed method using the healthy core samples to build the same four classifiers achieved significantly better classification accuracy (see Table 11). These results show that the use of a subset of the HCs from different datasets has the potential to support the generalization of classification models.

**Table 11.**
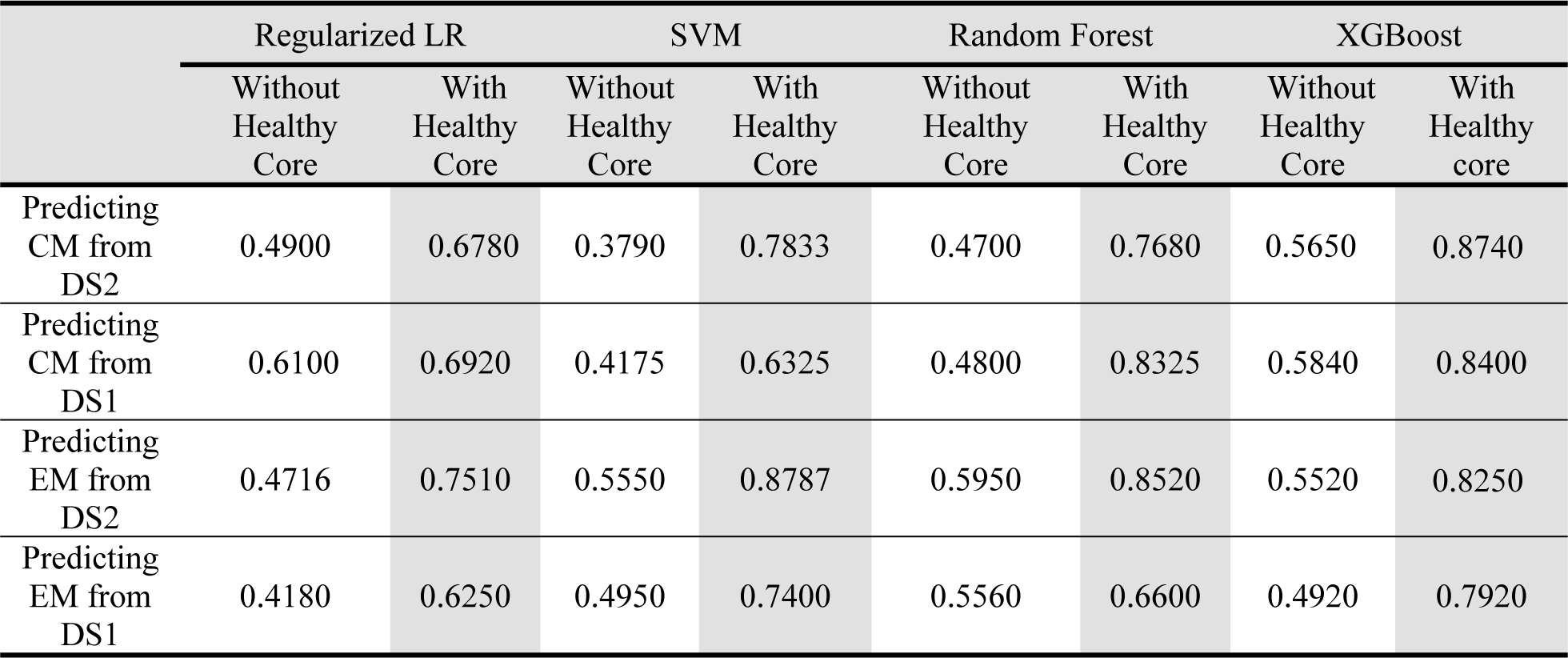
Accuracy of four models with and without Healthy Core for CM and EM Classification

### 3.5. Discussions on Healthy Core

In Section 3.1, we conducted statistical analysis to confirm the dataset discrepancy between DS1 and DS2. For illustration, here we adopted t-Distributed Stochastic Neighbor Embedding (t-SNE), a technique for dimensionality reduction (e.g., based on principal component analysis) that is particularly well-suited for the visualization of high-dimensional datasets to demonstrate the impact of a healthy core. For details of t-SNE, interested readers are referred to [Van der Maaten 2008].

As seen in Figure 3A and Figure 3C, combined HCs do not have separating (thus predictive) pattern compared to CM from either DS1 or DS2. On the other hand, the selected HCs cores have representative similar patterns, and healthy core (see Figure 3B and Figure 3D) can be well clustered in the t-SNE plot.

**Figure 3.**
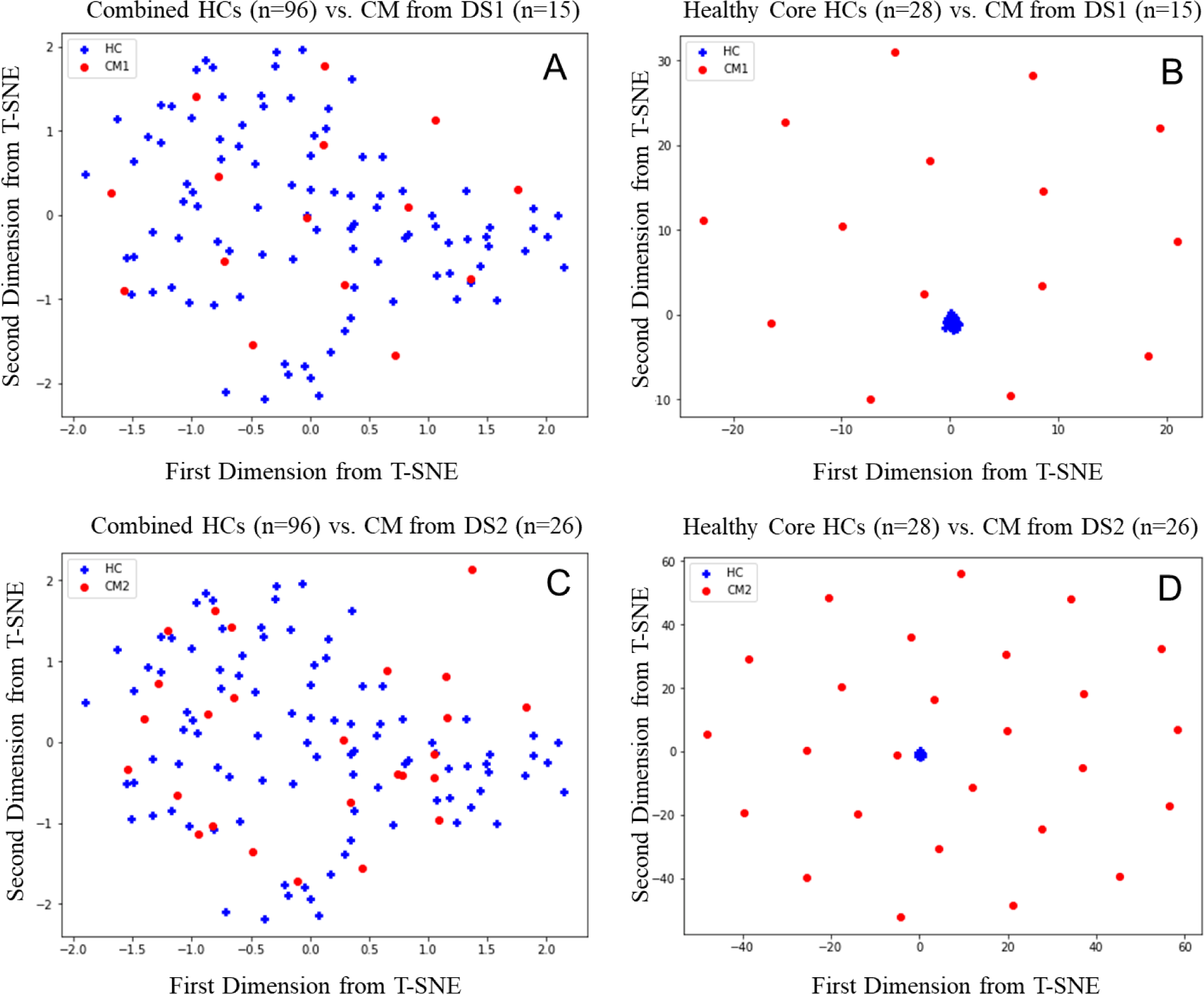
t-SNE plot of DS 1 and DS2 for CM on the first and second principal component. (A) combined HCs (n=96) vs. CM from DS1 (n=15). (B) Healthy Core (n=28) vs. CM from DS1 (n=15). (C) combined HCs (n=96) vs. CM from DS2 (n=26). (D) Healthy Core (n=28) vs. CM from DS2 (n=26).

Similar trends were observed for HC vs EM. Figure 4A demonstrated that the combined HCs contain mixed patterns with noise. However, 28 healthy cores identified by the proposed methods show compact representative patterns and are well separable from EM.

**Figure 4.**
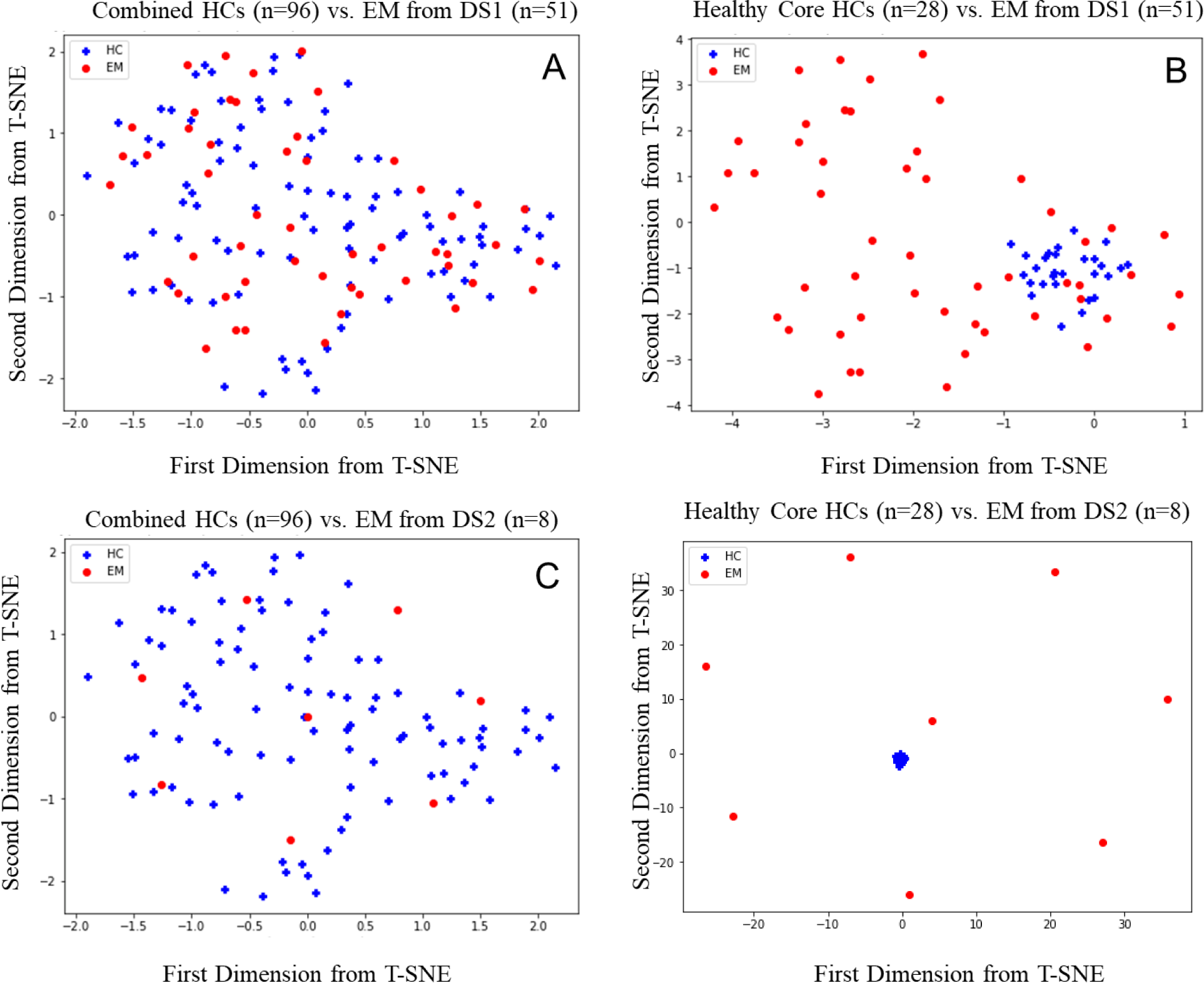
t-SNE plot of DS 1 and DS2 for EM on the first and second principal component. (A) combined HCs (n=96) vs. EM from DS1 (n=51). (B) healthy core (n=28) vs. EM from DS1 (n=51). (C) combined HCs (n=96) vs. EM from DS2 (n=8). (D) healthy core (n=28) vs. EM from DS2 (n=8).

## 4. Discussion and Conclusion

The main finding of this study is that utilization of a healthy core, i.e. a homogeneous dataset derived from a cohort of healthy controls, improves the performance of classification models for EM and CM. The use of a healthy core helps to overcome the deterioration in model performance that is otherwise seen when applying the migraine classification model to independent datasets collected from patients enrolled at different institutions and imaged on different scanners. Depending on the modeling approach, the inclusion of a healthy core resulted in classification accuracy as high as 88% for EM and CM.

The use of independent training and testing datasets is a substantial strength of the studies reported herein. Many prior publications (including some of our own) reported the accuracy of classification models for migraine train and test the model using the same dataset, with strategies such as the “leave-one-out” methodology[Schwedt, T.J. et al. 2015, Chong, C.D. et al., 2017, Lee M.J. et al., 2019, Zhang, Q. et al., 2018, Yang, H. et al., 2018, Chong, C.D., 2021]. Although this type of approach is valid and well-accepted, it likely leads to overestimating the performance of the model if it was used in a completely new and previously unseen dataset. Because independent training and testing datasets were used in the experiments described herein, the reported classification accuracies should be considered as more conservative estimates of model performance.

Useful classification models need to have high performance when tested on data that are collected from new patient populations and using collection techniques that might differ slightly from those used to collect the data that were included for model training (i.e. cross dataset accuracy). For example, a brain MRI-based classification model should still have high performance when tested on data from patients imaged at a different medical center and using different MR scanners. The experiments reported in this manuscript purposefully introduce this type of heterogeneity, with participants being enrolled from two different medical centers in two different regions of the United States. Results demonstrate the expected reduction in accuracy of the migraine classification models when they have been tested on independent datasets that include heterogeneity typical of multicenter imaging experiments. The experiments without the healthy core that are reported herein demonstrate this: EM and CM classification accuracy was higher in the single dataset experiments (CM accuracy 65%-75%, EM accuracy 61%-70%) compared to the cross-dataset experiments (CM accuracy 57%-59%, EM accuracy 58%-60%). The introduction of a healthy core helped to overcome this deteriorating model performance and provided much higher classification accuracies for EM and CM.

When performing research that includes “healthy controls” there is an assumption that the healthy control cohort is homogeneous. However, even when stringent eligibility criteria are applied in an attempt to make the healthy control group as healthy as possible, there will always be identifiable and unidentifiable heterogeneity within the healthy control group due to differences in demographics, prior life experiences, physical and mental well-being, underlying genetic heterogeneity, the existence of diseases that have not yet manifest, and other sources of heterogeneity. When establishing “normal values” for certain types of tests, such as blood test results, for example, the use of very large sample sizes can mostly overcome this issue of heterogeneity within the healthy control cohort. However, available sample sizes are smaller for establishing normal values for brain MRI, a diagnostic test that is time and cost-intensive. The “healthy core” method described in this manuscript can help overcome this challenge. For example, with the use of the healthy core and XGBoost, the cross dataset classification accuracy improved to 84%-87% for CM and to 79%-83% for EM.

When using the healthy core, the classification accuracies for migraine are comparable, if not somewhat higher, than those reported previously in the literature. This is so even though independent training and testing sets were used. For example, prior studies using brain imaging structural data reported classification accuracies of 67%-86% for differentiating migraine from healthy controls [Schwedt, T.J. et al. 2015], while those using functional MRI data reported accuracies of 73%-86% [Chong, C. D. et al. 2017; Yang, H et al. 2018; Lee, M. J. et al. 2019]. Classification models including multimodality imaging data, combining structural and functional measures, have provided accuracies of 83%-84 [Zhang, Q. et al. 2016; Gaw, N. et al. 2018]. Classification accuracies for EM were generally lower than for CM in the experiments reported herein. This is consistent with expectations since CM is the more severe disease state, prior brain imaging studies have demonstrated that headache frequency has a positive correlation with the amplitude of structural and functional brain changes, and prior classification models have had similar findings [Valfrè, W. et al. 2008; Schmitz, N. et al. 2008; Maleki, N. at al. 2012; Schwedt, T.J. et al. 2015; Gaw, N. et al. 2018; Magon, S. et al. 2019; Lai, K. L. et al. 2020; Mu, J. et al. 2020].

Study Limitations: We assumed heterogeneity amongst the healthy control cohort, and this assumption served as justification for developing a healthy core. However, a limitation of this study is that we are only partially able to describe the heterogeneity using the non-imaging data that were collected, such as the medical center from which a participant was enrolled, their sex, and age. Future studies should more deeply phenotype the healthy control subjects in search of sources of heterogeneity that might associate with differences in brain structure. Also, it should be noted that imaging data used in the analyses reported herein were also used in prior research on classification models for migraine [Schwedt, T.J. et al. 2015; Gaw, N. et al. 2018]. Future studies will continue to validate the migraine classification models reported herein and test the value of using a healthy core when developing new classification models.

In conclusion, the utilization of a healthy core can increase the accuracy and generalizability of brain imaging-based classification models for EM and CM. Inclusion of a healthy core addresses intrinsic heterogeneity that exists within a healthy control cohort and in multicenter studies, even when stringent participant eligibility criteria and methodology are used.

## Data Availability

Data from the study sponsored by the United States Department of Defense (DOD) will be made available through the Federal Interagency Traumatic Brain Injury Research (FITBIR) Informatics System in accordance with the rules and regulations of the DOD. Patient consent for the NIH-sponsored study and for the Mayo-funded study did not include a data sharing agreement.

## Appendix

The proposed method, a unified framework of GFK+MMD

The Maximum Mean Discrepancy (MMD) is a single statistic to measure the closeness between two distributions ℙ and ℚ from two datasets.

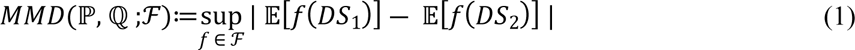

where ℱ is a set containing all continuous functions [Gretton, A et al. 2012]

The Geodesic Flow Kernel [Gong, B et al. 2012], also known as GFK, has utilized Grassmann manifolds and enabled the construction of physically meaningful new representations between two datasets. GFK explores several feature representations between two datasets and generates a number of transformations that characterize changes in geometric and statistical properties from one dataset to another dataset. Since this approach explores several statistically meaningful feature representations between two datasets, the advantage of several representations by GFK can be used for capturing data variabilities instead of relying on a single snapshot of each dataset. MMD is utilized as the tool to compare those feature representations.

**Algorithm 1:** a proposed algorithm for Healthy Core

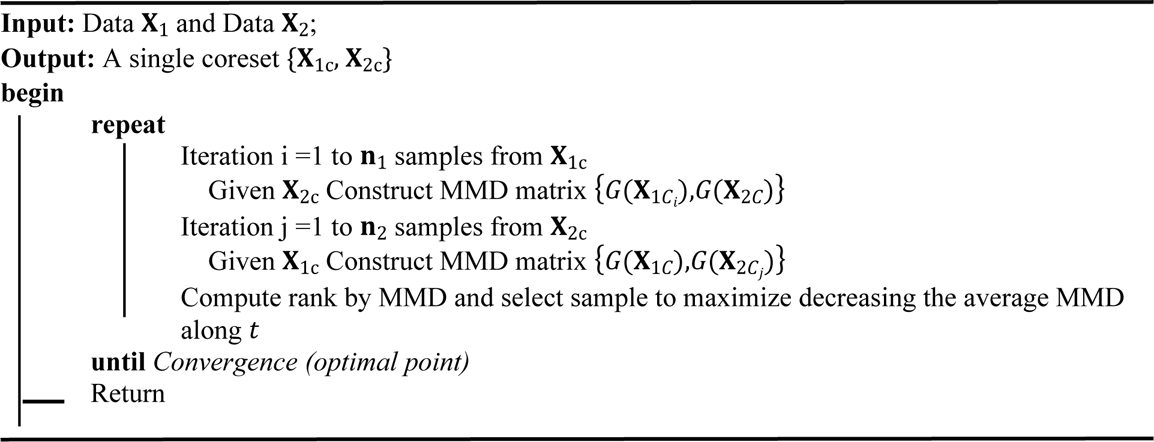

